# Precision Prostate Cancer Screening with a Polygenic Risk Score

**DOI:** 10.1101/2020.08.23.20180570

**Authors:** Tõnis Tasa, Mikk Puustusmaa, Neeme Tõnisson, Berit Kolk, Peeter Padrik

## Abstract

Prostate cancer (PC) is the second-most common type of cancer and the fifth-leading cause of cancer-related death in men worldwide. Genome-wide association studies have identified numerous genetic variants (SNPs) independently associated with PC. The effects of such SNPs can be combined into a single polygenic risk score (PRS). Stratification of men according to PRS could be applied in secondary prevention. We aimed to construct a PRS model and to develop a pipeline for personalized prostate cancer screening.

Previously published PRS models for predicting the risk of prostate cancer were collected from the literature. These were validated on the Estonian Biobank (EGC) consisting of a total of 16,390 quality-controlled genotypes with 262 prevalent and 428 incident PC cases and on 209 634 samples in the UK Biobank with 3254 prevalent cases and 6959 incident cases. The best performing model was selected based on the AUC in prevalent data and independently validated in both incident datasets. Using Estonian PC background information, we performed absolute risk simulations and developed individual risk-based clinical follow-up recommendations.

The best-performing PRS included 121 SNPs. The C-index of the Cox regression model associating PC status with PRS was 0.641 (SE = 0.015) with a hazard ratio of 1.65 (95% confidence interval 1.51 – 1.81) on the incident EGC dataset. The model is able to identify individuals with more than a 3-fold risk increase. The risk of an average 45-year old could be attained by individuals between the ages of 41 and 52. A 41-year old male on the 95th percentile has the same risk as an average 45-year old but by age 55, he has attained the same genetic risk as an average 68-year-old.

PRS is a powerful predictor of prostate cancer risk that can be combined with current non-invasive practices of PC screening. We have developed PRS-based recommendations for personalized PSA testing. Our approach is easily adaptable to other nationalities by using population-specific background data of other genetically similar populations.

## Introduction

Prostate cancer (PC) is the second-most common type of cancer that caused more than 350,000 deaths in 2018 (1). In EU countries, PC accounted for more than 10% of all male cancer deaths (1). Mortality rates of PC have recently decreased in most Western nations; considered to be partly due to a widely adopted aggressive prostate PC screening policy (2).

Many methods with different performances are available for PC detection: the prostate-specific antigen (PSA) level, digital rectal examination of the prostate gland, prostate volume measurement, magnetic resonance imaging, trans-rectal ultrasonography, and prostate biopsy (3). The European Association of Urology currently supports the use of family history, magnetic resonance imaging, prostate volume, and digital rectal examination to serve as tools in PC triage and baseline PSA testing at the age of 45 to individualize screening intervals (4).

Genetics provides an enhancement to the risk stratification toolbox (5) as PC risk has significant heritability – 57% (95% confidence interval [CI] 51%-63%) (6). Clinically important high-penetrance variants of BRCA1, BRCA2, ATM, CHEK2 genes are rare, but their carriers have a 2–4 fold increase in the risk of developing PC in their lifetime than the general population. Testing of these gene variants should be done in healthy men with a reported family history or according to other pre-specified criteria (7-9). Importantly, most PC genetic predisposition is polygenic.

Genetic testing for prostate cancer is driven by the prospect of informing early detection and individual screening strategies (10, 11). In the last decade, more than 160 PC risk-associated single nucleotide polymorphisms (SNPs) have been identified through genome-wide association studies that explain around 30% of total risk (12-15). In contrast to high-penetrance variants, these PC risk-associated SNPs are common and with small effects that can be combined into a stronger cumulative effect via a polygenic risk score (PRS) with predictive utility (15-19). The PRS predicts prostate cancer risk better than family history but since it is only moderately heritable even in first-degree relatives then it needs individual estimation (20). Polygenic predisposition can be used in combination with existing techniques such as PSA to personalize early detection strategies with potential reductions in over-diagnoses and false positives (21-23).

This study aims to evaluate the risk prediction performance of several published prostate cancer PRS models and to assess the best-performing model use as a risk stratification approach in the context of Estonia. Concretely, we aim to combine polygenic risks with a low-cost PSA based screening strategy.

## Methods

### Biobank participant data

We used validation data from two population biobanks: the Estonian Biobank of the Estonian Genome Center at the University of Tartu (EGC) and the UK Biobank (UKBB). Quality controlled samples were divided into prevalent and incident datasets. The prevalent dataset included PC cases diagnosed before Biobank recruitment with 5 times as many controls without the diagnosis. Incident data included cases diagnosed in any of the linked databases after recruitment to the Biobank and all controls not included in the prevalent dataset. Prevalent datasets were used for identifying the best candidate model and the EGC incident dataset was used to obtain an independent PRS effect estimate on PC status.

### Participant data of Estonian Genome Center

PC cases and controls in retrospective data of EGC were defined by ICD-10 code (C61) status derived from questionnaires filled at recruitment of the gene donors and from linked data from Estonian Cancer Registry (data until 2013), National Health Insurance Fund (data until the end of 2018) and Causes of Death Registry (data until 2017 May).

All EGC samples were genotyped in Core Genotyping Lab of Institute of Genomics, University of Tartu, using Illumina GSAMD-24v1–0 arrays. Individuals were excluded if the total variant call-rate was < 95% or sex defined based on X chromosome heterozygosity did not match declared sex. Variants were filtered by call-rate < 95%, HWE p-value < 1e-4 (autosomal variants only) and minor allele frequency < 1%. Variant positions were updated to b37 and all variants were changed to the TOP strand (https://www.well.ox.ac.uk/~wrayner/strand/). Phasing was done using Eagle (v. 2.3) software (24) and imputation with Beagle (v. 28Sep18.793) (25) using the Population-specific imputation reference of 2297 WGS samples (26).

### Participant data of UK Biobank

This study used genotypes from the UK Biobank cohort (version v3, obtained 07.11.2019) and made available to Antegenes under application reference number 53602. The data was collected, genotyped using either the UK BiLEVE or Affymetrix UK Biobank Axiom Array. PC cases in the UK Biobank cohort were retrieved by the status of ICD-10 code C61. We additionally included cases with self-reported UK Biobank code “1044”.

Quality control steps and in detail methods applied in imputation data preparation have been described by the UKBB and made available at http://www.ukbiobank.ac.uk/wpcontent/uploads/2014/04/UKBiobank_genotyping_QC_documentation-web.pdf. We applied additional quality controls on autosomal chromosomes. First, we removed all variants with allele frequencies outside 0.1% and 99.9%, genotyping call rate < 0.1, imputation (INFO) score < 0.4 and Hardy-Weinberg equilibrium p-value < 1E-6. Sample quality control filters were based on several pre-defined UK Biobank filters. We removed samples with excessive heterozygosity, individuals with sex chromosome aneuploidy, and excess relatives (> 10). Additionally, we only kept individuals for whom the submitted gender matched the inferred gender and genotyping missingness rate was below 5%.

### Model selection from candidate risk models

We searched the literature for PRSs in the public domain. The requirements for inclusion in the candidate set were availability of the chromosomal location, reference and alternative allele, minor allele frequency, and an estimator for the effect size either as odds ratio (OR) or its logarithm (log-OR) specified for each individual genetic variant. In cases of iterative model developments on the same underlying base data, we retained chronologically newer ones. The search was performed with Google Scholar and PubMed web search engines by working through a list of articles using the search [“Polygenic risk score” or “genetic risk score” and “prostate cancer”], and then manually checking the results for the inclusion criteria. We additionally pruned the PRS from multi-allelic, non-autosomal, non-retrievable variants based on bioinformatics re-analysis with Illumina GSA-24v1 genotypes and non-overlapping variants between EGC and UKBB data.

PRSs were calculated as 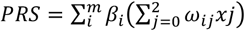 where *ω_i_* is the probability of observing genotype *j*, *where j*∈*(0,1,2)* for the i-th SNP; *m* is the number of SNPs; *β_i_* is the effect size of the i-th SNP estimated in the PRS. The mean and standard deviation of PRS in the cohort were extracted to standardize individual risk scores to Gaussian. We tested the assumption of normality with the mean of 1000 Shapiro-Wilks test replications on a random subsample of 1000 standardized PRS values.

Next, we evaluated the relationship between PC status and standardized PRS in the two prevalent datasets with a logistic regression model to estimate the logistic regression-based odds ratio per 1 standard deviation of PRS (*OR_sd_*), its p-value, Akaike information criteria (AIC) and Area Under the ROC Curve (AUC). The logistic regression model was compared to the null model using the likelihood ratio test and to estimate the Nagelkerke and McFadden pseudo-R^2^. We selected the candidate model with the highest AUC to independently assess risk stratification in the incident datasets.

### Independent performance evaluation of a polygenic risk score model

The main aim of the analyses in the incident datasets was to derive a primary risk stratification estimate, hazard ratio per 1 unit of standardized PRS (HR*sd)*, using a right-censored and left-truncated Cox-regression survival model. The start of time interval was defined as the age of recruitment; follow-up time was set as the time of diagnosis for cases and at the time of last health data linkage for controls. Scaled PRS was used as the only independent variable of PC diagnosis status. 95% confidence intervals were created using the standard error of the log-hazard ratio. We also assessed the goodness-of-fit of the survival model using the Harrell’s C-index and the likelihood ratio test.

Further, we evaluated the concordance between theoretical hazard ratio estimates derived with the continuous per unit PRS (HRsd) estimate and the hazard ratio estimates inferred empirically from data. For this, we binned the individuals by PRS to 5%-percentiles and estimated the empiric hazard ratio of PC directly between those classified in each bin and those within the 40–60 PRS percentile. Theoretically estimated hazard ratio estimates assume a multiplicative effect of the mean in a PRS bin on the unit based hazard ratio. This relationship between HRsd and the expected mean in the truncated Gaussian PRS distribution is expressed as 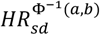, where the exponent is the mean of a truncated Gaussian distribution between two percentiles *a* and *b* (bounded between 0 and 1, a<b), and 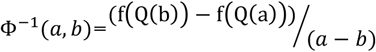 where *Q(b)* is the Gaussian quantile function on a percentile *b* and *f(Q(b))* is the Gaussian probability density function value at a quantile function value. We compared the two approaches by using the Spearman correlation coefficient and the proportion of distribution-based 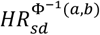 estimates in empirical confidence intervals.

### Absolute risk estimation

Individual *τ*-year (eg. 10-year) absolute risk calculations are based on the risk model developed by Pal Choudhury et al. (27). Individual absolute risks are estimated for currently *a*-year old individuals in the presence of known risk factors *(Z)* and their relative log hazard-ratio parameters (*β*). 95% uncertainty intervals for the hazard ratio were derived using the standard error and z-statistic 95% quantiles CI_HR_ = exp(*β* ±1.96*se(HR)), where se(HR) is the standard error of the log-hazard ratio (HR) estimate. Risk factors have a multiplicative effect on the baseline hazard function. The model specifies the next *τ*-year absolute risk for a currently *a*-year old individual as

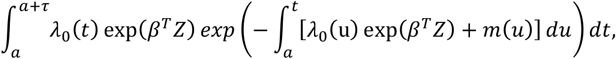

where *m(t)* is age-specific mortality rate function and *λ_0_(t))* is the baseline-hazard function, *t* ≥ *T* and *T* is the time to onset of the disease. The baseline-hazard function is derived from marginal age-specific PC incidence rates (*λ*_*m*_*(t)*) and distribution of risk factors *Z* in the general population *(F(z))*.

This absolute risk model allows disease background data from any country. In this analysis, we used Estonian background information. We calculated average cumulative risks using data from the National Institute of Health Development of Estonia (28) that provides population average disease rates in 5-year age groups, and group sample sizes from Statistics Estonia for 2013–2016. Per person-year incidence rates for each age group were calculated as *IR*=*X_t_/N_t_* for data from 2013–2016, where X_t_ is the number of first-time cases at age *t* and N*_t_* is the number of exposed individuals in this age group. Final per-year incidences were averaged over time range 2013–2016. With incidence rates calculated based on binned groups, we assumed the incidence rates to stay constant throughout the aggregation period.

Age-and sex-specific mortality data for the year 2016 was retrieved from World Health Organization (29) and competing mortality rates were constructed by subtracting yearly age- and sex-specific disease mortality rates from general mortality rates. Prostate cancer mortality estimates were extracted from the Global Cancer Observatory (30).

We applied this model to estimate absolute risks for individuals in the 1^st^, 10^th^, 25^th^, 50^th^, 75^th^, 90^th^ and 99^th^ PRS quantiles, eg. an individual on the 50^th^ percentile would have a standardized PRS of 0. Confidence intervals for the absolute risk are estimated with the upper and lower confidence intervals of the continuous per unit log-hazard ratio. Similarly, we used the absolute risk model to estimate lifetime risks (between ages 0 and 85) for the individuals in the same risk percentiles.

### PRS based risk-stratification and individual screening recommendations

Next, we simulate PRS risk separation in the Estonian population background context. Our analysis first established the 10-year risk of a 45-year old male with a population average of PRS (“average male”) as the reference for the level of risk deeming continuous monitoring. Here, we assessed the differences in ages where individuals in various PRS risk percentiles attain 1 to 3-fold risk increases of risk compared to the 10-year risk of an average male. Based on these analyses, we developed recommendations for a PC monitoring program that develops a routine for prostate specific antigen (PSA) testing and test-based follow up recommendations. The PRS risk stratification uses both relative risks, fold difference of 10-year risks compared to a genetically average individual of the same age and sex, and also her absolute 10-year risk.

## Results

### Polygenic risk score re-validation in population cohort datasets

In the EGC cohort, we retained a total of 16,390 quality-controlled male samples. All samples were divided into prevalent and incident datasets. Altogether, 262 cases were prevalent (ie. disease diagnosis before Biobank recruitment) and combined with 1310 controls. All other cases and controls were included in the incident dataset, combining for a total of 13,390 controls and 428 cases.

The larger UKBB dataset contained 209 634 samples that passed the quality controls. In UKBB, we identified 3254 prevalent cases and 6959 incident cases that were complemented with 16 245 controls and 183 176 controls, respectively.

Altogether, we identified 5 PRS models that were to be evaluated. Normality assumption of the standardized PRS was only violated with the model PC15 that we were not able to successfully replicate (Shapiro-Wilks test p-values in EGC data PC1 = 0.47, PC2 = 0.24, PC3 = 0.08, PC4 = 0.37, PC15 = 7.8e-15). Table 1 highlights that the best model is PC3 based on AUC, ORsd, AIC, and pseudo-R^2^ metrics in both EGC and UKBB data. The Corresponding AUC under the ROC curve (Figure 1) for the association between the PRS and PC diagnosis was 0.631 (SE = 0.04) in EGC and 0.632 (SE = 0.011) in UKBB.

**Table 1.** Comparison metrics of PC PRS models based on the prevalent Estonian Genome Center dataset.

|  |  | PC1 (31) | PC2 (16) | PC3 (12) | PC4 (32) | PC15* (19) |
| --- | --- | --- | --- | --- | --- | --- |
| Variants in original PRS |  | 103 | 25 | 147 | 93 | 48 |
| Variants included in analyses |  | 92 | 10 | 121 | 82 | 39 |
| Estonian<br>Genome<br>Center | AUC (SE) | 0.592<br>(0.044) | 0.566<br>(0.041) | 0.631<br>(0.042) | 0.585<br>(0.045) | 0.541<br>(0.044) |
|  | OR<sub>sd</sub> ((SE [log<br>OR<sub>sd</sub>])) | 1.42 (0.07) | 1.33 (0.07) | 1.61 (0.07) | 1.40 (0.07) | 0.90 (0.07) |
|  | AIC | 1394.1 | 1404.3 | 1371.5 | 1396.7 | 1418 |
|  | McFadden /<br>Nagelkerke<br>Pseudo-R<sup>2</sup> | 1.9% / 2.8% | 1.1% / 1.7% | 3.5% / 5.1% | 1.7% / 2.5% | 0.17% /<br>0.26% |
| UK Biobank | AUC (SE) | 0.677<br>(0.011) | 0.600<br>(0.012) | 0.685<br>(0.011) | 0.665<br>(0.011) | 0.513<br>(0.012) |
|  | OR<sub>sd</sub> ((SE [log<br>OR<sub>sd</sub>])) | 1.95 (0.02) | 1.43 (0.02) | 1.98 (0.02) | 1.83 (0.02) | 0.94 (0.02) |
|  | AIC | 16 481 | 17 259 | 16 342 | 16 637 | 17 578 |
|  | McFadden /<br>Nagelkerke<br>Pseudo-R<sup>2</sup> | 6.3% / 9.2% | 1.8% / 2.8% | 7.1% /<br>10.4% | 5.4% / 8.0% | 0.06% /<br>0.09% |

\*PC15 – Likely problems with adapting allele directionalities reported in the original analysis to our study.

**Figure 1.**
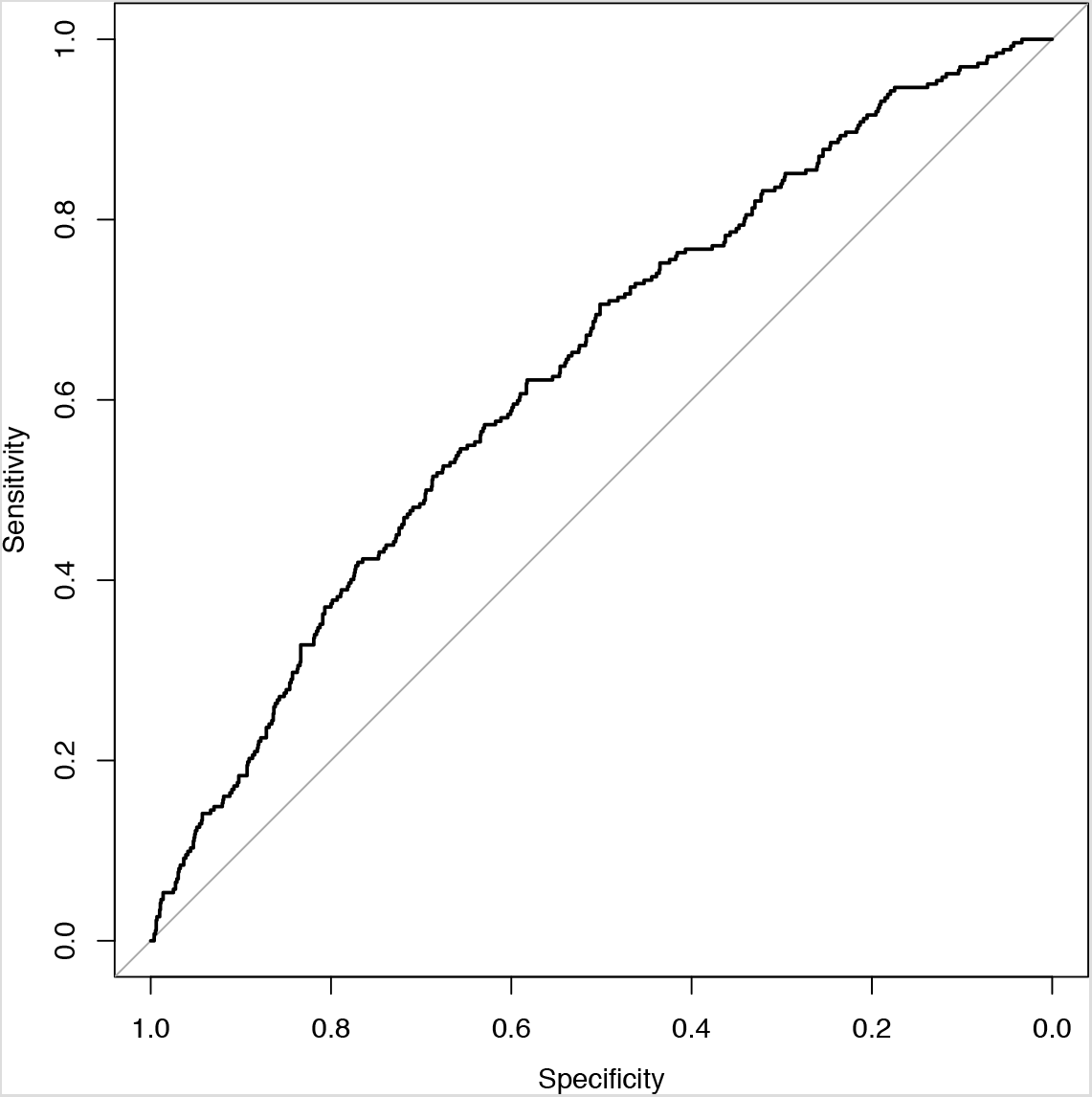
ROC plot of PC cases and controls in prevalent Estonian Genome Center dataset.

Next, we evaluated the performance of the best performing PC3 model in the independent incident datasets with the main aim of estimating the hazard ratio per unit of PRS. Table 2 presents the performance estimation metrics. Hazard ratio per 1 unit of standard deviation (HR_sd_) in model PC3 was *1.65* with standard error (log *(HR)*) = 0.05). The concordance index (C-index) of the survival model testing the relationship between PRS and PC diagnosis status in the incident EGC dataset was 0.641 (se = 0.015) and 0.654 (SE = 0.003) in UKBB.

**Table 2.**
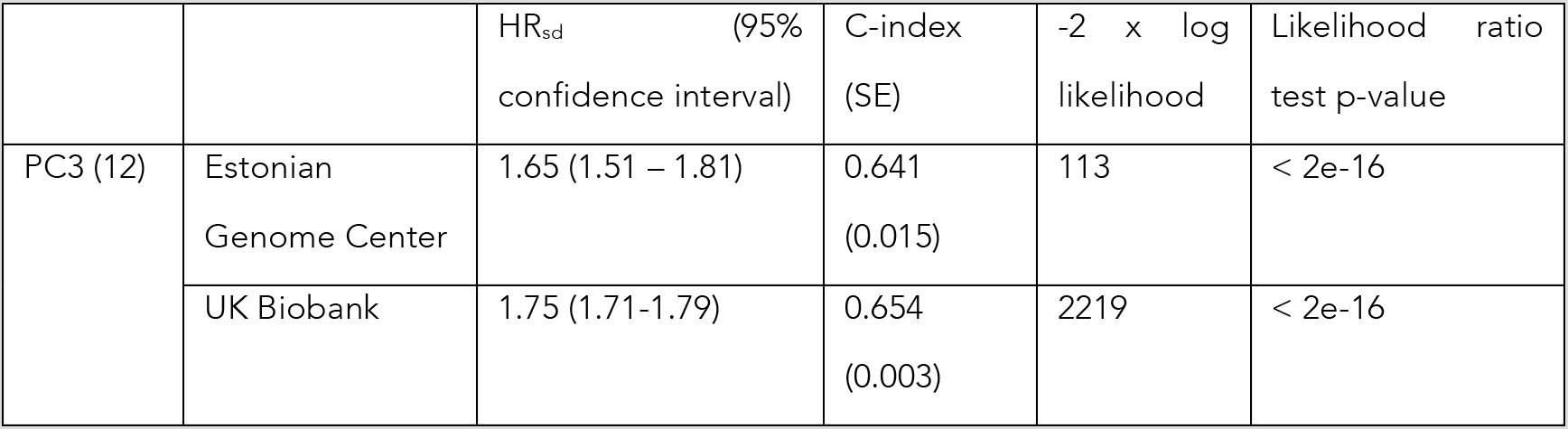
Performance metrics of the Cox regression model on the disease status and PC3 based polygenic risk scores calculated in the incident dataset.

|  |  | HR<sub>sd</sub> (95% confidence interval) | C-index (SE) | -2 x log likelihood | Likelihood ratio test p-value |
| --- | --- | --- | --- | --- | --- |
| PC3 (12) | Estonian Genome Center | 1.65 (1.51 – 1.81) | 0.641 (0.015) | 113 | < 2e-16 |
|  | UK Biobank | 1.75 (1.71-1.79) | 0.654 (0.003) | 2219 | < 2e-16 |

Figure 2 highlights the trend of hazard ratio estimates compared to individuals in the 40–60 percentile of PRS. In panel A, the theoretical hazard ratio matched empirical estimate’s confidence intervals in all comparisons. Alternatively, in panel B with UKBB data, the Spearman correlation coefficient between the empiric and theoretical hazard ratio estimates is 0.936, indicating very strong association, and the much narrower confidence bands match in 15 out of 16 cases.

**Figure 2.**
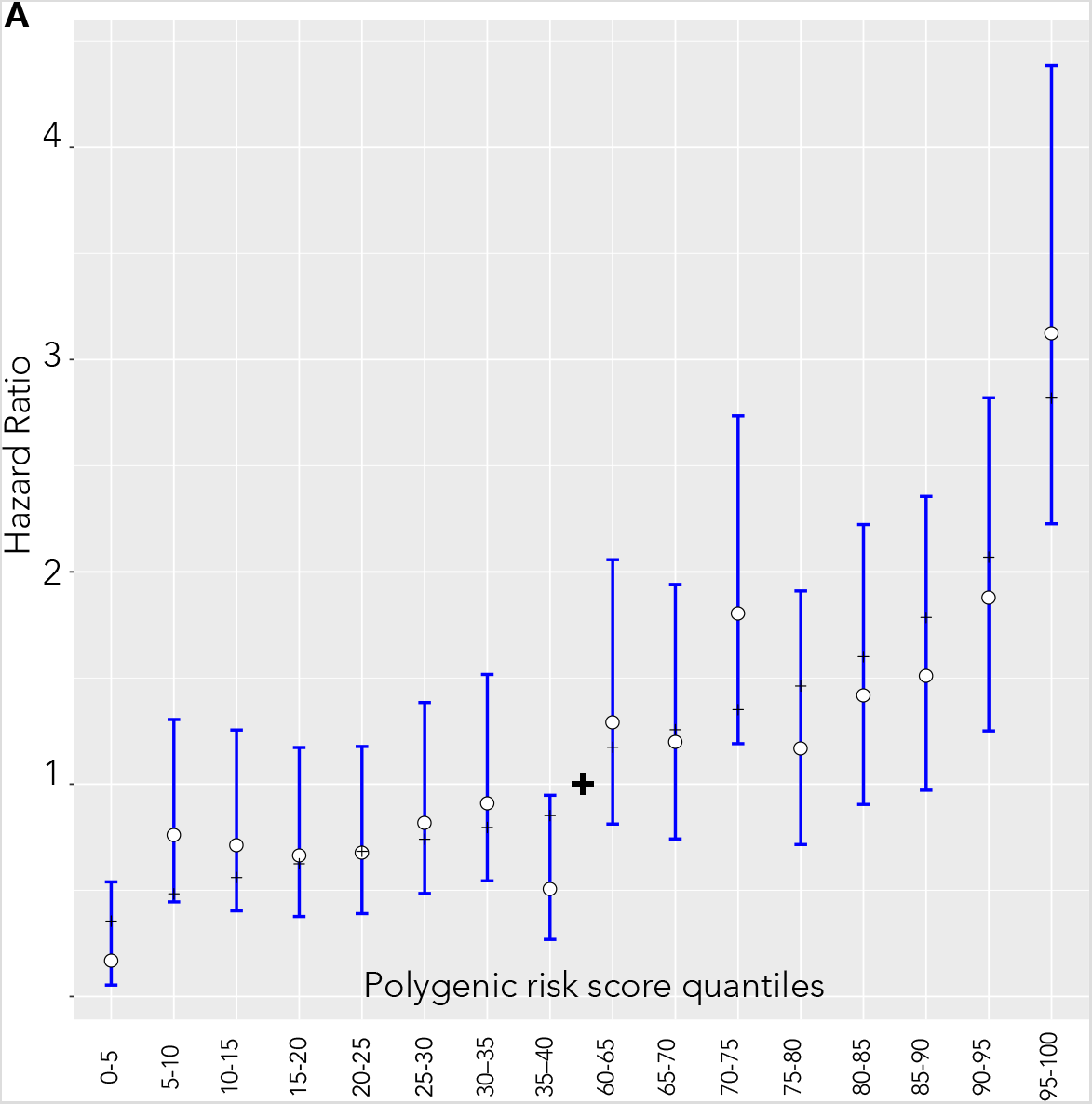
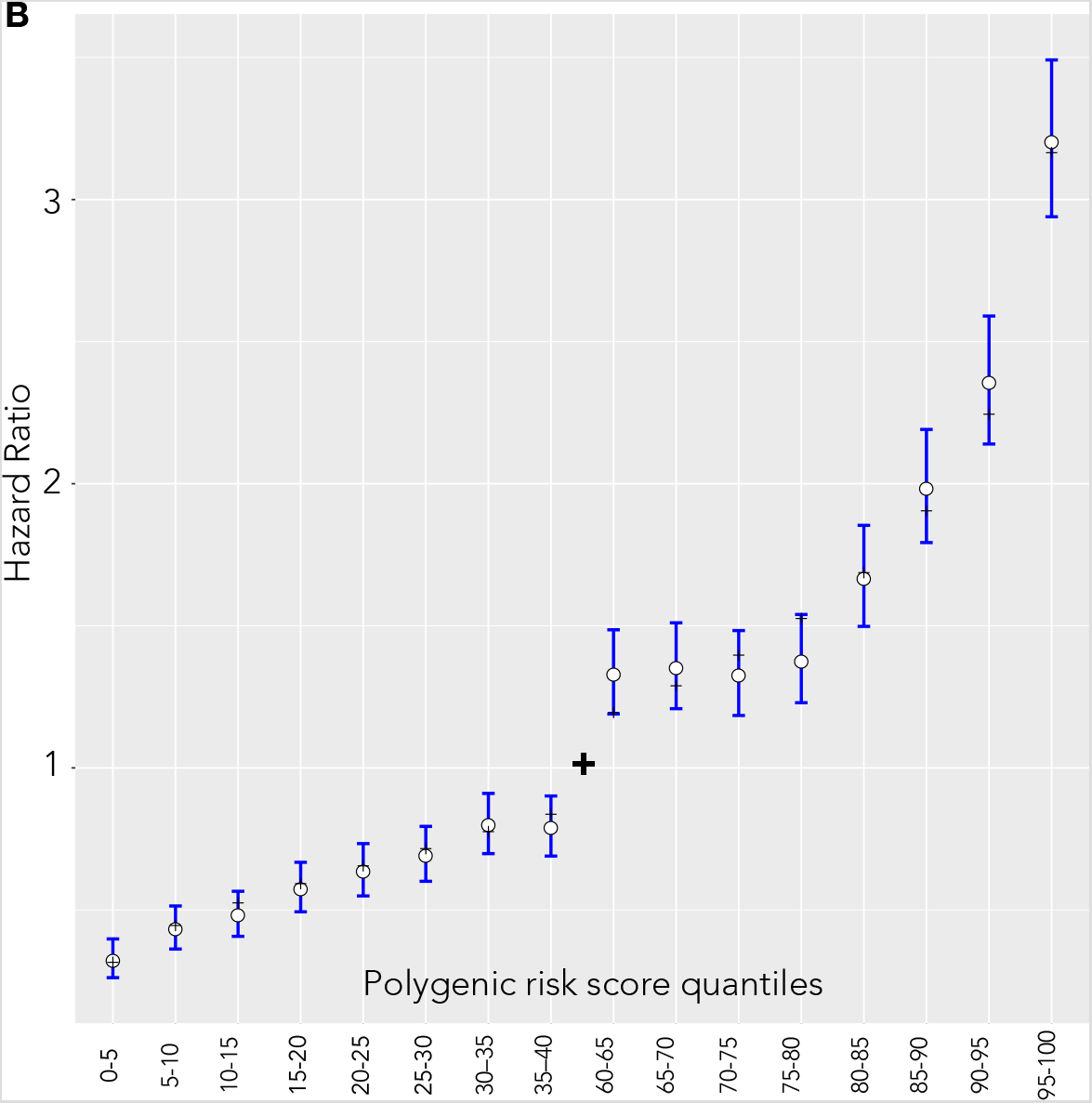
Hazard ratio estimates between quantiles 40–60 of the PC3 PRS and categorized 5% bins in the incident dataset. White dots and blue lines represent empirically estimated hazard ratio estimates and corresponding confidence intervals. Black dashes represent the theoretical hazard ratio for the 5%-quantile bins derived from the hazard ratio of per unit PRS. (A) Estonian Genome Center (B) UK Biobank.

### Polygenic risk score in prostate cancer screening stratification

We used a model by Choudhury et al. to derive individual 10-year risks (27) and specified *F(z)* as the distribution of PRS estimates in the whole EGC cohort. The log-hazard ratio (β) is based on the estimate of the log-hazard ratio in the PC3 model of the incident EGC dataset. Age-specific PC incidence and competing mortality rates provided the background for PC incidences in the Estonian population.

Prostate cancer 10-year risk for those in the 1^st^ percentile of PRS among 45-year old Estonian males was 0.18% (0.14% – 0.22%) and 1.81% (1.53% – 2.11%) for those in the 99^th^ percentile, and 2.19% (1.72% – 2.78%) and 20.56% (17.4% – 24.1%) respectively for the same percentiles at age 70. The relative risks between the most extreme percentiles are therefore around 9.3-fold. Competing risk accounted cumulative risks reach 30.7% (26.7% – 35.0%) by age 85 for those in the 99^th^ percentile but remain at 3.69%- (2.89% – 4.69%) for those in the lowest 1^st^ percentile (Figure 3).

**Figure 3.**
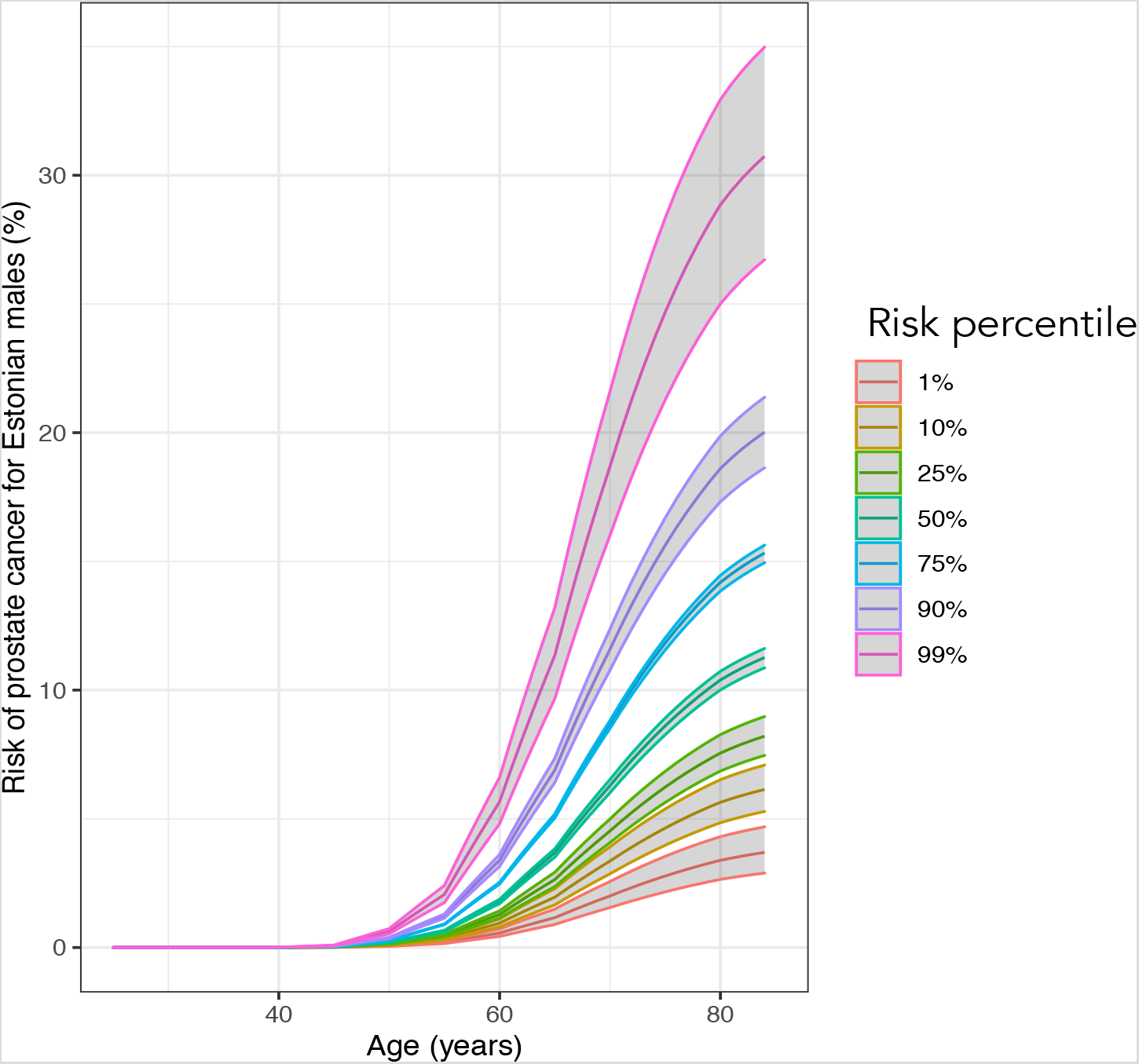
Cumulative risks (%) of PC between ages 20 to 85 in various risk percentiles

A genetically average 45-year old male has a 10-year absolute risk of around 0.57%. PT3 model can identify 41-year-old males in the 99^th^ percentile of PRS that have a larger risk than the average risk of 45-year-olds. At the same time, males in the 1^st^ percentile (and below) attain this risk by their 52^nd^ birthday. In effect, individual men could be at the average 45-year-old male risk anywhere between ages 41 and 52. Similarly, males above 92^nd^ of PRS percentile have a more than 2-fold and around > 1% of males attain a 3-fold risk compared to those at average risk. PC incidence increases rapidly with age. We observe that individuals in the 1^st^ percentile attain the 3-fold risk of the average 45-year old male already by age 61 (Figure 4).

**Figure 4.**
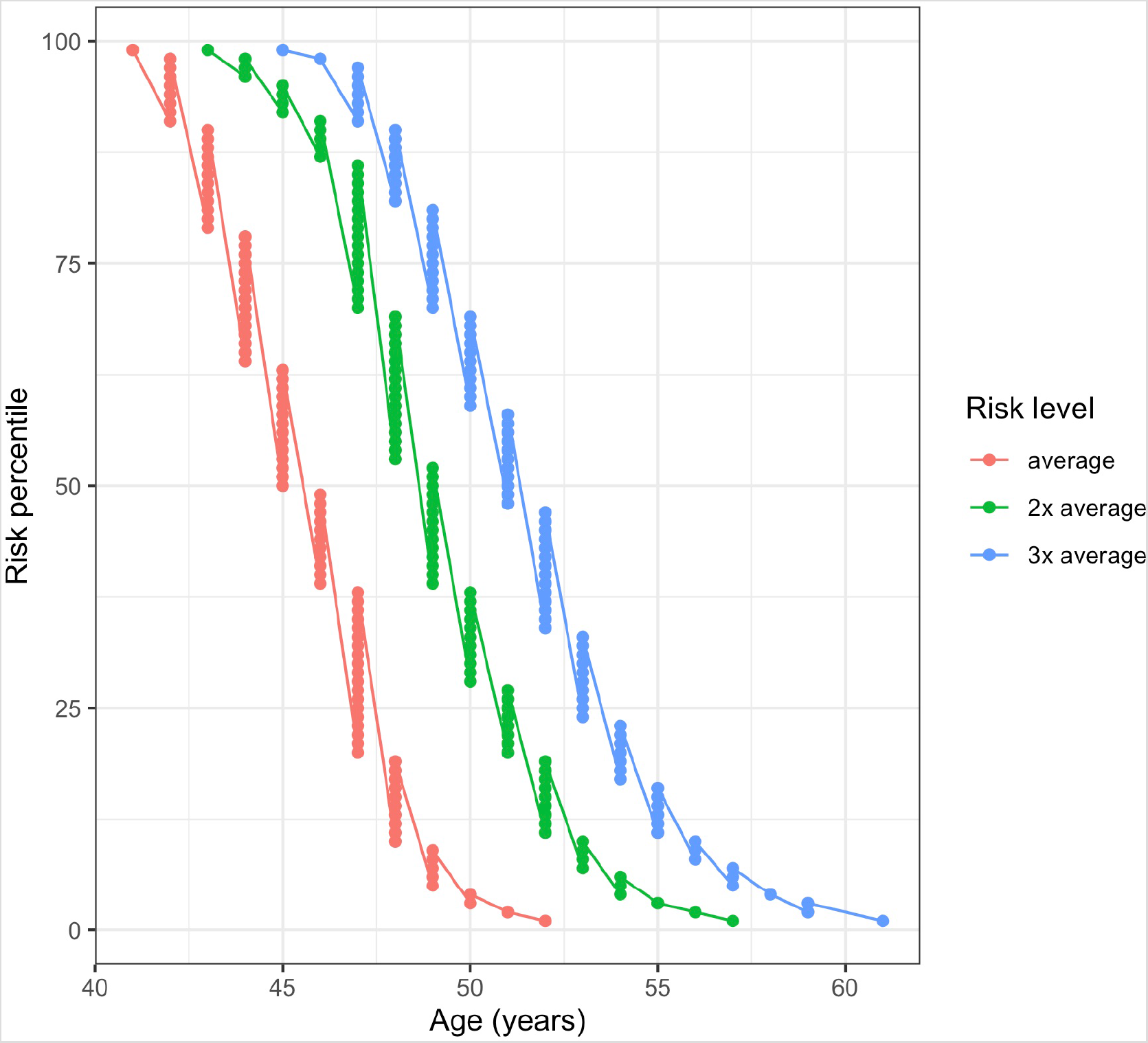
Ages when Estonian females in different risk percentiles attain 1–3 fold multiples of 10-year risk compared to 45-year old males with population average PRS (Risk level: “average”).

A 41-year old male on the 95th percentile has the same risk as an average 45-year old but by age 55, he has attained the same genetic risk as an average 68-year-old.

PRS provides a basis for personalized screening recommendations that are based on relative risks compared to an individual of the same nationality, age, and sex, and also the estimated absolute risks. PC risks start increasing rapidly from age 45 so genetic risk-based recommendations presented below are based on increasing surveillance activities based on attaining the risk multiples of genetically average 45-year old. PC screening recommendations with PSA testing based on guidelines from the EAU are also accompanied by general guidelines for reducing the risk of PC. PSA testing and digital rectal examination if possible, are recommended for early detection of PC. According to the international guidelines, it is necessary to provide an individualized risk-adjusted strategy for the early detection of PC in well-informed men with a good general status and predictable life expectancy of at least 10–15 years.

1. Relative risk is less than 1.5-fold higher than average

a. PSA testing from age 45.

i. PSA test value at the age of 45-60: < 1 ng/ml – further PSA testing in 8-year intervals;
ii. PSA test value at the age of 45-60: ≥ 1 ng/ml – further PSA testing in 2-year intervals.
iii. PSA test value over 60: < 2 ng/ml – further PSA testing in 8-year intervals;
iv. PSA test value over 60: ≥ 2 ng/ml – further PSA testing in 2-year intervals.

2. The relative risk increased between 1.5 and 3-fold.

a. PC screening with two-year intervals starting at the age of attaining a 10-year risk of a genetically average 45^*^ year old.
b. If possible, digital rectal palpation of the prostate by a doctor for screening.

3. The relative risk increased more than 3-fold

a. PC screening with two-year intervals starting from age 40 and a one-year interval from age 50.
b. If possible, digital rectal palpation of the prostate by a doctor for screening.

^*^*If the recommended age is below the individual’s current age, then recommend current age*

## Discussion

In this study, we qualified and re-validated the performance of a pruned 121 PRS model by Schumacher *et al* (12). Our model performance results were consistent with previous literature. We applied this model to design a novel absolute risk-based PC screening strategy based on Estonian screening information and background data. The model identifies more than 3-fold differences in risk. In the Estonian population, the individuals can attain the risk of an average 45-year old between ages 41 and 52. However, relative genetic effects increasingly bring about differences in absolute risk attainment with growing age. We combined information from PC PRS with current clinical guidelines about PSA testing intervals and reference values.

Studies reported here have previously reported their performance (12, 16, 19, 31, 32). Our validation analyses did not account for other variables besides PRS. Al Olama *et al*. reported a strong association between PRS and PC risk, with an OR per standard deviation of PRS of 1.74 (95% CI 1.70–1.78). After adjusting for age and PC family history, men with PRS in the highest 10% of the distribution were 2.31 (95% CI 2.09–2.56) times as likely to develop PC as those between the 25th-75th percentiles of PRS (16). Schumacher *et al*. estimated that men with PRS in the top 1% are at roughly 5.7-fold higher PC risk than average and those in the top 10% with nearly 2.7-fold higher. They reported an odds ratio of 1.86 per unit of PRS (95% CI 1.83–1.89) after adjusting for batch effects and principal components (12). We were not able to successfully replicate the PRS reported by Seibert *et al*. (2018) due to likely ambiguities in reported reference and effect alleles. This PRS was reported to identify men in the top 2% of the score with an almost 3-fold greater relative risk for aggressive prostate cancer compared to average (19). Previously reported metrics are in general agreement with the ones reported here.

Knowledge of greater genetic risk predisposition allows for instituting primary prevention activities (33). However, there is no final consensus on the utility of prostate cancer screening (11, 34-37). Some extended follow-up programs indicate that the mortality reduction remains unchanged (38), even as screening results in increased diagnoses and detection of more localized disease cases (37). False positives and overtreatment of the benign disease need to be balanced against potential benefits (39). Others report that men undergoing regular PSA testing with around 20% lower chance of dying from PC (40) and a 30% lower chance of developing the metastatic disease (41) compared with men not performing PSA testing. Importantly, current approaches with non-invasive and inexpensive PSA screening can be potentially leveraged with other types of data (23).

Combining PRS with PSA has shown the potential for increasing the performance of stand-alone screening (18). Chen et al. found an AUC of 88.8% (95% CI 88.6–89.1) for PSA combined with the 7 SNP PRS compared to 70.1% (95% CI 69.6–70.7) for PSA alone. Further increases to 96.7% (95% CI 96.5–96.9) were observed when a larger number of PC susceptibility variants were included. The prospective Stockholm 3 aimed to develop a model to identify high-risk PC with better test characteristics than provided by PSA screening alone (42). Performing regular PSA monitoring in combination with PRS provides a pathway to timely diagnostic activities.

The foremost challenge is the communication of the PRS that motivates the uptake of requisite preventative measures. Knowledge of personal genetic risk has not been convincingly associated with implemented behavioral or lifestyle changes. Thoughtful feedback should, therefore, guide uptakes of therapeutic recommendations that are intrinsically linked with evidence-based intervention recommendations (43). Our approach is easily adaptable to nationalities other than Estonia by using population background information data of other genetically similar populations. Similarly, the clinical screening recommendations can be adapted to locality specific screening environments as long as we can infer the absolute risk of the average male in that locality.

In conclusion, we have used a PRS based model to develop a novel model for PC screening. Our PRS identifies individuals at more than 3-fold risk and identified that the effect of genetics on absolute risk attainment becomes increasingly pronounced after age 40. The genetic risk-based recommendations can be applied prospectively by individuals and also by institutions aiming to link genetic testing with current prostate cancer screening activities.

## Data Availability

Individual level genotype and phenotype data from Estonian Biobank or UK Biobank can not be explicitly shared. The UK Biobank Resource was used under Application Reference Number 53602. New users can request access to UK Biobank from http://www.ukbiobank.ac.uk/resources/. Similarly, Estonian Biobank data is available by request. Researchers interested in Estonian Biobank can request the access here: https://www.geenivaramu.ee/en/access-biobank

https://www.geenivaramu.ee/en/access-biobank

## Acknowledgments

This research has been conducted using the UK Biobank Resource under Application Reference Number 53602 and with support by EIT Health the Digital Sandbox program.

As in the related paper on the development of breast cancer polygenic risks scores, our appreciation goes to everybody from the Estonian Genome Center actively involved in this project. Specifically, we would like to than Kristi Läll for recommendations on the manuscript and proof-reading, prof. Krista Fischer for feedback in the study planning phase, Reidar Anderson, Merli Saare and Reedik Mägi for supporting the data exchange and running the analyses in EGC, and our friends Viljo Soo and Mari Nelis for great work in the laboratory. We fully appreciate the efforts of prof. Lili Milani, prof. Andres Metspalu and prof. Tõnu Esko for their trust and support in developing this initiative.

Also, we would not have been able to perform any analyses without the computation resources provided by Sander Kuusements and Ivar Koppel from the HPC Center of the University of Tartu.

## Ethical approval

### Estonian Genome Center

All human research was approved by the Research Ethics Committee of the University of Tartu, and conducted according to the Declaration of Helsinki and Human Research Act. All participants provided written informed consent to participate in the Estonian Biobank.

### UK Biobank

The UK Biobank study was approved by the North West Multi-Centre Research Ethics Committee (UK Biobank reference: 16/NW/0274). All participants provided written informed consent to participate in the UK Biobank study.

